# Improving estimates of pertussis burden in Ontario, Canada 2010-2017 by combining validation and capture-recapture methodologies

**DOI:** 10.1101/2022.08.05.22278478

**Authors:** Shilo H. McBurney, Jeffrey C. Kwong, Kevin A. Brown, Frank Rudzicz, Andrew Wilton, Natasha S. Crowcroft

**Affiliations:** Dalla Lana School of Public Health, University of Toronto, Toronto, Ontario, Canada; Public Health Ontario, Toronto, Ontario, Canada; Department of Laboratory Medicine and Pathobiology, University of Toronto, Toronto, Ontario, Canada; ICES, Toronto, Ontario, Canada; Department of Family & Community Medicine, University of Toronto, Toronto, Ontario, Canada; Department of Computer Science, University of Toronto, Toronto, Ontario, Canada; International Centre for Surgical Safety, Li Ka Shing Knowledge Institute, St. Michael’s Hospital, Toronto, Ontario, Canada; Vector Institute for Artificial Intelligence, Toronto, Ontario, Canada; Immunization, Vaccines and Biologicals, World Health Organization, Geneva, Switzerland

## Abstract

An underestimation of pertussis burden has impeded understanding of transmission and disallows effective policy and prevention to be prioritized and enacted. Capture-recapture analyses can improve burden estimates; however, uncertainty remains around incorporating health administrative data due to accuracy limitations. The aim of this study is to explore the impact of pertussis case definitions and data accuracy on capture-recapture estimates. We used a dataset from March 7, 2010 to December 31, 2017 comprised of pertussis case report, laboratory, and health administrative data. We compared Chao capture-recapture abundance estimates using prevalence, incidence, and adjusted false positive case definitions. The latter was developed by removing the proportion of false positive physician billing code-only case episodes after validation. We calculated sensitivity by dividing the number of observed cases by abundance. Abundance estimates demonstrated that a high proportion of cases were missed by all sources. Under the primary analysis, the highest sensitivity of 78.5% (95% CI 76.2-80.9%) for those less than one year of age was obtained using all sources after adjusting for false positives, which dropped to 43.1% (95% CI 42.4-43.8%) for those one year of age or older. Most code-only episodes were false positives (91.0%), leading to considerably lower abundance estimates and improvements in laboratory testing and case report sensitivity using this definition. Accuracy limitations can be accounted for in capture-recapture analyses using different case definitions and adjustment. The latter enhanced the validity of estimates, furthering the utility of capture-recapture methods to epidemiological research. Findings demonstrated that all sources consistently fail to detect pertussis cases. This is differential by age, suggesting ascertainment and testing bias. Results demonstrate the value of incorporating real time health administrative data into public health surveillance if accuracy limitations can be addressed.

## Introduction

Pertussis remains one of the most common vaccine-preventable diseases in Canada [1]. Despite being a reportable disease, an underestimation of cases and deaths has impeded understanding of transmission. Ascertainment bias is a key concern, which occurs when atypical cases are underdiagnosed including older individuals experiencing mild disease [2, 3]. This issue is worsened by testing bias, with younger, severe cases more likely to be tested, have a positive result, and be reported [4, 5]. Burden estimates vary regionally based on interactions between case definitions, the type of surveillance and data available, practitioner knowledge, immunization programs, and the extent of local transmission [4, 6]. Underestimation may be attributed to failing to consider pertussis diagnostically, atypical presentations, infrequent diagnostic testing, suboptimal test accuracy, lack of uniformity in case definitions, and reporting issues [2, 5]. In combination with complicated epidemiological characteristics, pertussis surveillance is consequently challenging [4, 7]. However, monitoring burden is essential for informing and assessing the impact of immunization programs and policy [4, 6-9].

To improve pertussis burden estimates, one strategy is to supplement surveillance data with health administrative data [10]. When several sources are available, capture-recapture analyses can be used to better estimate burden [11]. This analytic approach has been recently used to assess completeness of contact-tracing for Ebola and detection of Covid-19 infections [12, 13]. For pertussis, capture-recapture has been used to estimate the number of deaths in England and the number of cases in Ontario [10, 14]. The latter estimated that 21-73% of cases have been missed using combined surveillance, laboratory, and health administrative data [10]. However, considerable uncertainty in estimation remained around the validity of using the latter, and particularly Ontario Health Insurance Plan (OHIP) physician billing diagnostic codes. The aim of this study is to explore the impact of using different pertussis case definitions and adjusting for data accuracy on capture-recapture results, with the goal of enhancing the utility of this method for improving burden estimates to inform public health surveillance, prevention, and policy.

## Methods

The University of Toronto’s Health Sciences Research Ethics Board (37885) and the Public Health Ontario (PHO) Ethics Review Board (2019-006.02) approved this study. Data were linked and analyzed at ICES (formerly the Institute of Clinical Evaluative Sciences) using unique encoded identifiers.

### Data sources

We obtained Public Health Information System (iPHIS) and PHO Laboratory Information System (Labware) data from a PHO linked dataset previously used to improve estimates of pertussis burden in Ontario [10]. iPHIS data contained confirmed, probable, and “does not meet” (DNM) pertussis case reports. We considered reports greater than 365 days apart a new case, with data available from April 1, 2006 to March 31, 2015. When duplicates occurred in the same year, we gave priority to the highest level of confirmation. Labware data included positive, indeterminate, and negative pertussis laboratory tests, with cases defined as at least one positive result by PCR or culture. We counted positive results more than 90 days apart as a new case, and data were available from December 7, 2009 to March 31, 2015.

We updated the PHO dataset until March 31, 2018 and combined it with health administrative data from December 1, 2009 from three databases held at ICES: the Canadian Institute for Health Information (CIHI) Discharge Abstract Database (DAD); the CIHI National Ambulatory Care Reporting System (NACRS); and the Ontario Health Insurance Plan (OHIP) Claims Database (Fig 1). Collected data included ICD-10 codes A37.0 (whooping cough, *Bordetella pertussis)* and A37.9 (whooping cough, unspecified species) from hospitalizations and emergency room visits and OHIP diagnostic billing code 033 (whooping cough, *Bordetella pertussis*). We restricted OHIP claims to billings from homes, offices, and long-term care facilities. The Registered Persons Database (RPDB) was used to obtain data on patient sex, age, and date of death. We excluded health administrative data entries with same-day immunizations as they are unlikely to reflect a true pertussis case (Fig 1). We excluded all entries with no index date or a date of death prior to the index date, as well as participants with an invalid unique identifier or who were missing their sex or birth date (Fig 1).

**Fig 1.**
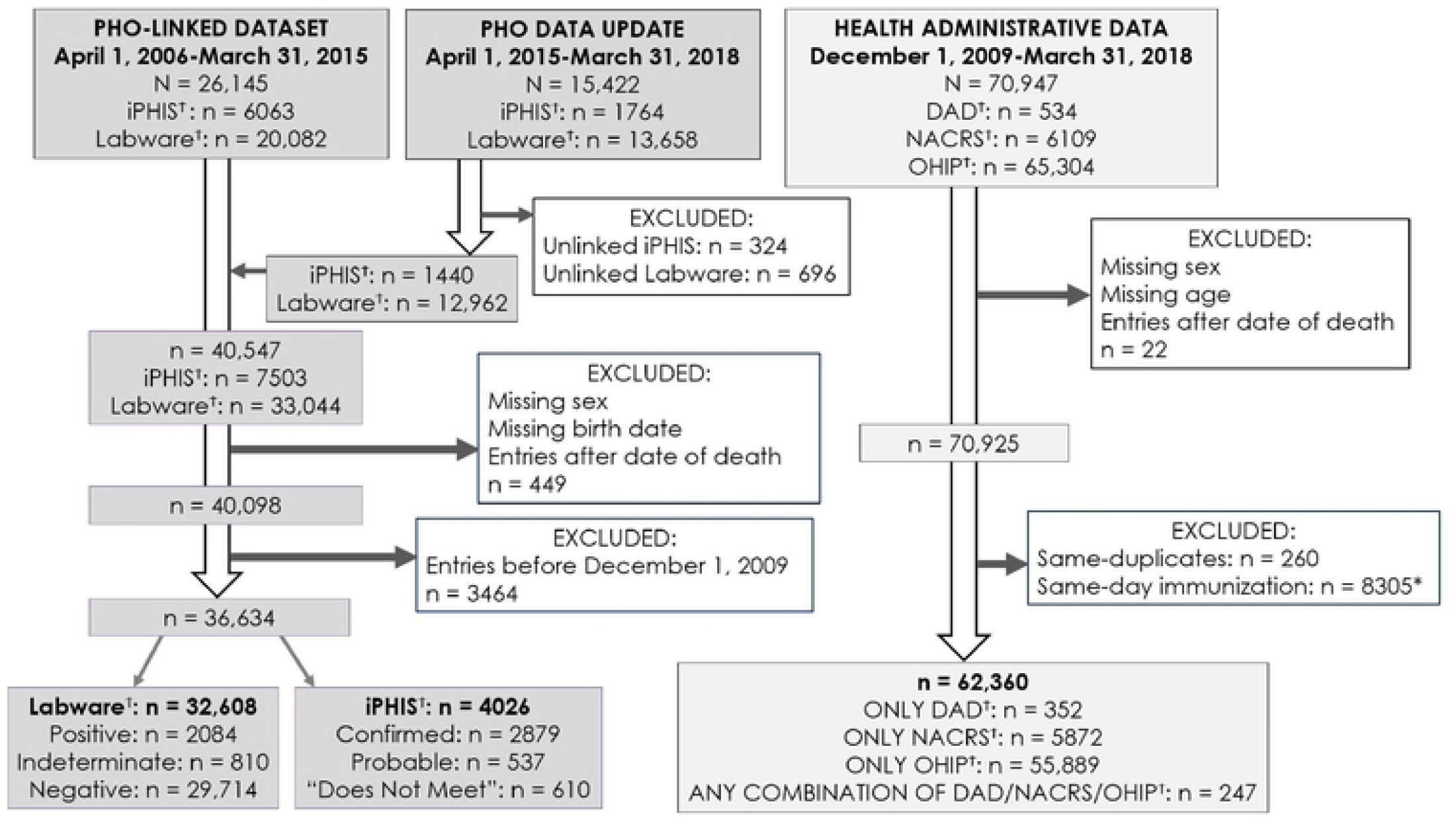
Data flow chart. ^Ϯ^OHIP = physician diagnostic billing codes, DAD = hospitalizations, NACRS = emergency room visits, iPHIS = reportable cases, Labware = laboratory tests, *using pertussis-containing immunization codes G840, G841, and G847 and general immunization codes G538 and G539.

### Case definitions and sensitivity analyses

We developed three case definitions to separately input into capture-recapture models – period prevalence, incidence with exclusions, and false-positive adjusted (Fig 2). Period prevalence permitted an individual to have one entry per data source over the study period. We did not apply a time limit to recapture in other sources, leading to the best recapture scenario. For incidence with exclusions, we incorporated data entries into episodes using 90-day rules and ruled out administrative data-only episodes with a negative pertussis laboratory test within 28 days of the episode start (Fig 2 and S1 Fig). This structure required recapture in other sources to occur within the 90 days before or after the respective episode. Finally, for the last definition we eliminated the proportion of false positive OHIP code-only episodes from the incidence with exclusions case definition. To do so, we validated these episodes to obtain a positive predictive value (PPV) and took a random sample based on the estimated proportion of true positives. We conducted validation using a previously developed methodology and cohort from the Electronic Medical Record Primary Care (EMRPC) database (S1 Appendix) [15, 16]. The PPV was estimated as 8.99%, leading to the removal of 41,062 OHIP-code only entries for the primary analysis (Fig 2).

**Fig 2.**
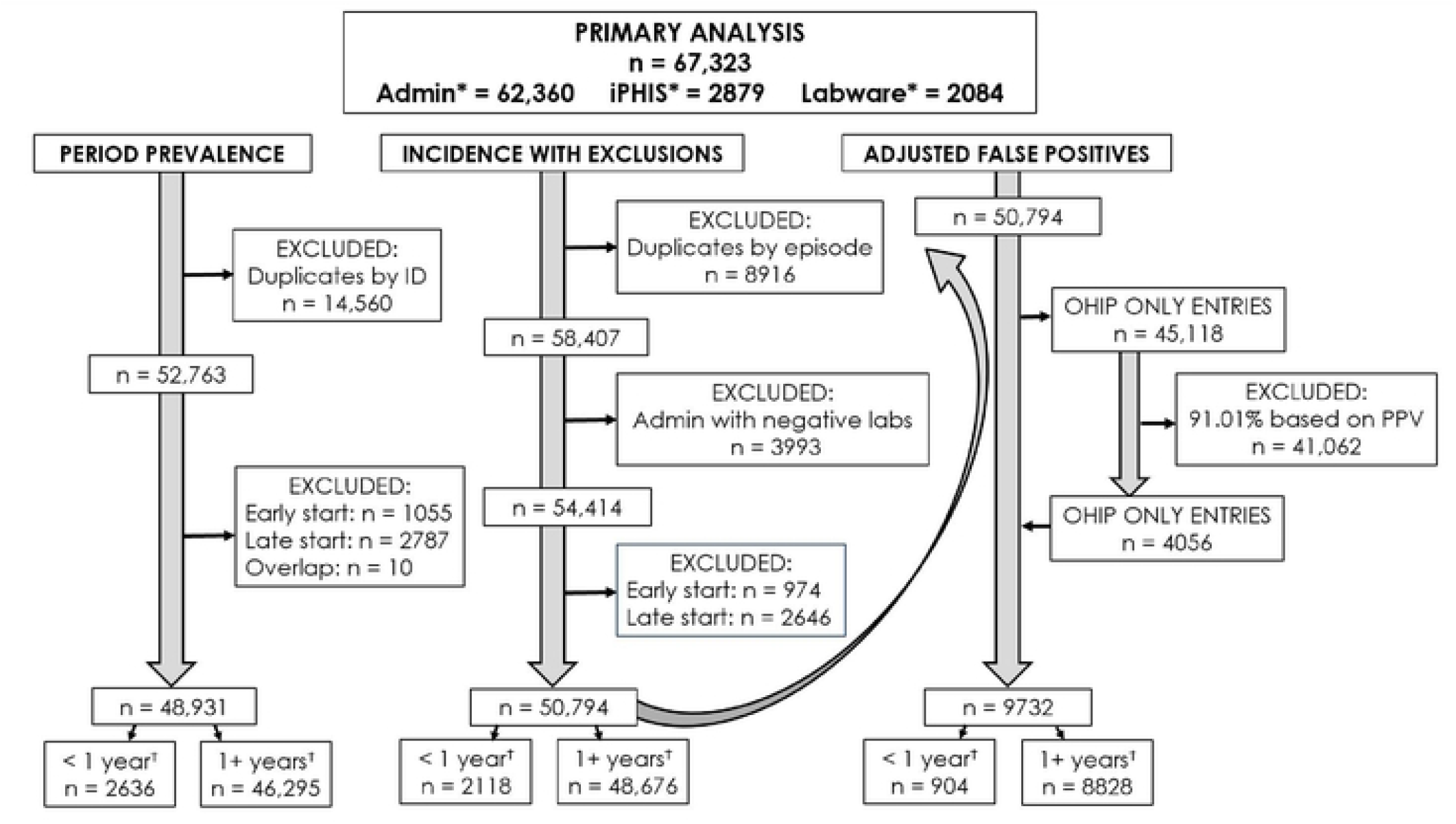
Flow diagram of case definitions. ^*^Admin= OHIP physician diagnostic billing codes, DAD hospitalizations, NACRS emergency room visits, iPHIS = reportable cases, Labware = laboratory tests, ^Ϯ^stratified by age groups, < 1 year of age and 1+ years of age.

To ensure that failure to recapture was not due to data missingness, we applied a study window based on data availability. We removed cases with a first date before March 7, 2010 and last date after December 31, 2017 to give a buffer of 90 days at each end to ensure an accurate episode start and end date, which we defined as the earliest and latest date for an episode in any data source. Finally, we applied three sensitivity analyses to the three data structures to explore further uncertainty in case definitions. This included incorporating iPHIS probable cases, iPHIS probable and DNM cases in addition to indeterminate laboratory tests, and excluding A37.9 codes with pertussis species unspecified. For the second sensitivity analysis, we ruled out episodes with a negative pertussis laboratory test within 28 days of the episode start if the episode only included administrative data or DNM entries.

### Capture-recapture analyses

Capture-recapture estimates abundance using the cases identified in each source and their overlap to calculate the number missed by all [17]. We assumed the population was closed and that temporality was present [10, 18]. We combined all administrative data into a single source at the outset, assuming and thereby accounting for dependency [11]. We assessed other dependencies by calculating the probability of being captured in one source given being in another [10]. Additionally, we evaluated random detection using Pearson’s chi-squared tests of the observed versus expected number of cases in a pair of sources [10]. Both were assessed under the prevalence structure to ensure the assumption of independence for these tests was not violated. We used these results to select a dependency structure in combination with theoretical considerations [19]. We evaluated heterogeneity by assessing the linearity of heterogeneity graphs [18].

We used models that accounted for a closed population, temporality, and heterogeneity if determined to be present. The latter was expected as milder, older cases of pertussis are less likely to be tested, have a positive result, and be reported to surveillance [4, 5]. We explicitly built the selected dependency structure into models by including two-way interaction coefficients for sources hypothesized to be dependent [11]. We chose Chao’s lower bound estimator for total sample size for the capture-recapture models, which additionally accounts for dependency [11]. We compared model estimates to those from Darroch models, which further correct for heterogeneity [20]. We rounded estimates of abundance to the nearest whole number. We considered results statistically significant at alpha ≤ 0.05 and we used the Rcapture package in R [18, 21].

### Estimated sensitivity

We calculated sensitivity by dividing the number of cases identified by each data source by the estimated abundance [10]. For the incidence and adjusted false positive case definitions, there was concern that correlation (clustering) would arise from having multiple cases for some individuals, impacting the sensitivity point estimate and variance [22, 23]. To address this, sensitivity was additionally calculated under both definitions by selecting a random episode per person to remove the effect of clustering. We then compared these results to those including multiple episodes, with little difference used as evidence that estimates were robust.

## Results

### Capture-recapture results

Under the prevalence case definition, all sources were dependent based on pair-wise probabilities and Pearson’s chi-squared test was highly significant (p < 0.00001), suggesting non-random detection. This provided support for the theorized dependency structure, and we used all two-way interaction terms to account for dependency between each source pair. To ensure comparability between case definitions, we used this structure for all capture-recapture models. A visual depiction of dependency is presented in Fig 3 using the degree of overlap between data sources under prevalence. Heterogeneity graphs lacked linearity, indicating heterogeneity was present and had to be accounted for in modelling (S2 Fig).

**Fig 3.**
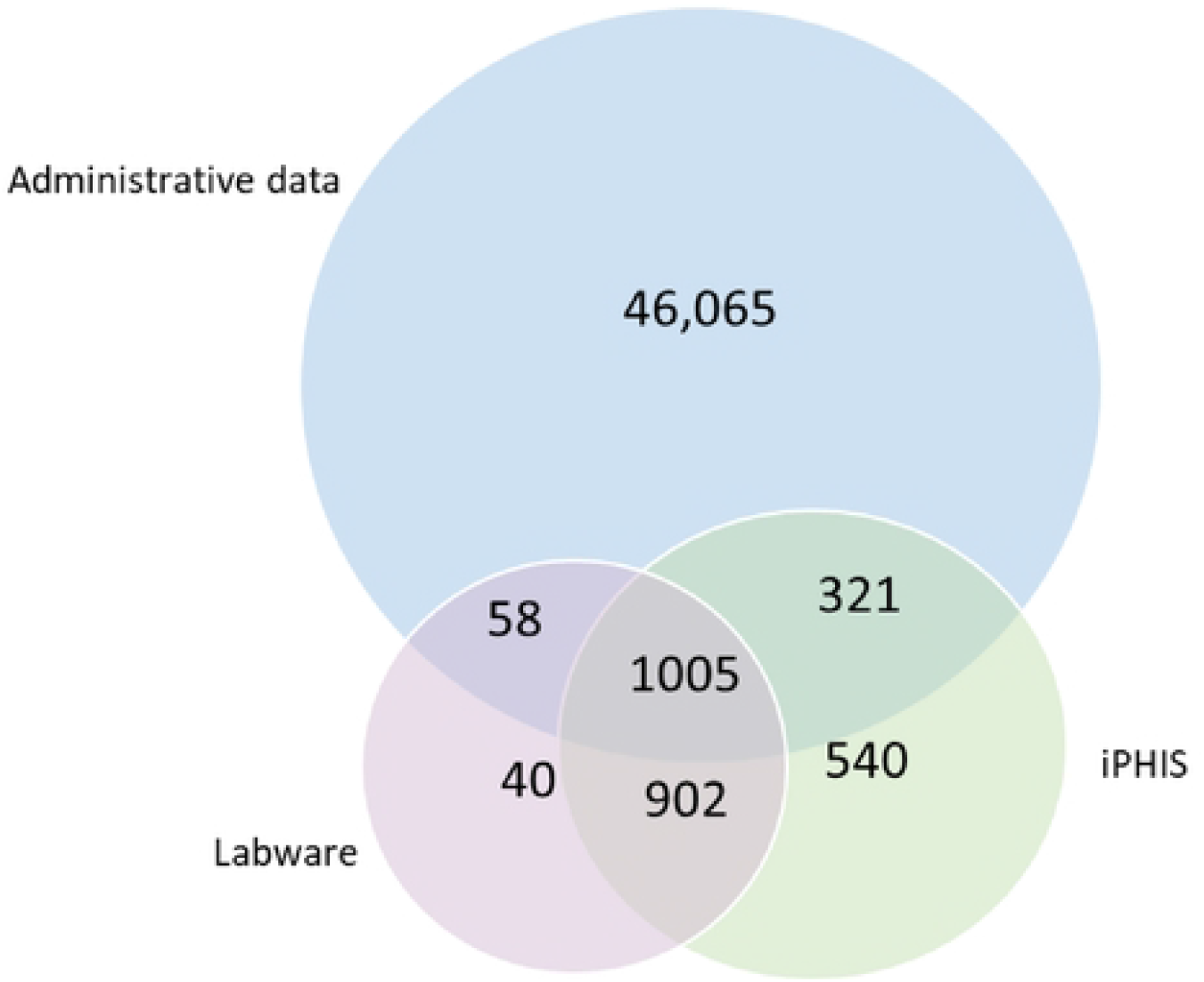
Example of overlap between data sources^Ϯ^, all age groups combined under the period prevalence primary analysis. ^Ϯ^Administrative data = OHIP physician diagnostic billing codes, DAD hospitalizations, NACRS emergency room visits, iPHIS = reportable cases, Labware = laboratory tests.

Estimated abundance was similar for those less than one year of age under the definitions for period prevalence and incidence with exclusions, at 3810 (95% CI 2932-5707) and 3078 (95% CI 2362-4609) respectively (Table 1). Abundance was considerably lower with less variability, as measured by the width of the 95% CIs, for the adjusted false positive definition at 1151 (95% CI 964-1538). Overall, the one year or older age group had more variable estimates. Abundance for this age group was again similar under prevalence and incidence, at 114,135 (95% CI 87,228-155,391) and 132,528 (95% CI 100,384-181,775). The false positive definition had substantially lower estimates and variability at 20,490 (95% CI 15,998-27,319). Darroch models produced identical abundance estimates in scenarios with more than one two-way interaction term.

**Table 1.** Capture-recapture model results with all two-way interactions between sources by analysis, age group, and case definition.

|  | Total observed pertussis cases | Chao model with all two-way interactions |  |  |
| --- | --- | --- | --- | --- |
|  |  | Estimated abundance | 95% CI | AIC, BIC* |
| PRIMARY ANALYSIS |  |  |  |  |
| < 1 year of age |  |  |  |  |
| Period prevalence | 2636 | 3810 | 2932-5707 | 56.9, 98.1 |
| Incidence with exclusions | 2118 | 3078 | 2362-4609 | 56.8, 96.4 |
| Adjusted false positives | 904 | 1151 | 964-1538 | 55.4, 89.1 |
| 1+ years of age |  |  |  |  |
| Period prevalence | 46,295 | 114,135 | 87,228-155,391 | 69.9, 131.1 |
| Incidence with exclusions | 48,676 | 132,528 | 100,384-181,775 | 70.1, 131.6 |
| Adjusted false positives | 8828 | 20,490 | 15,998-27,319 | 68.1, 117.7 |
| SENSITIVITY ANALYSIS 1† |  |  |  |  |
| < 1 year of age |  |  |  |  |
| Period prevalence | 2648 | 4030 | 3003-6240 | 57.3, 98.5 |
| Incidence with exclusions | 2133 | 3235 | 2418-4974 | 57.1, 96.8 |
| Adjusted false positives | 897 | 1163 | 963-1575 | 55.7, 89.3 |
| 1+ years of age |  |  |  |  |
| Period prevalence | 46,581 | 102,954 | 80,534-136,981 | 70.8, 132.1 |
| Incidence with exclusions | 49,015 | 119,684 | 92,638-160,724 | 71.1, 132.7 |
| Adjusted false positives | 9295 | 19,120 | 15,339-24,804 | 69.1, 119.0 |
| SENSITIVITY ANALYSIS 2† |  |  |  |  |
| < 1 year of age |  |  |  |  |
| Period prevalence | 2729 | 9197 | 6180-14,619 | 60.7, 102.1 |
| Incidence with exclusions | 2206 | 7883 | 5205-12,740 | 60.1, 100.0 |
| Adjusted false positives | 1007 | 2454 | 1759-3691 | 58.8, 93.2 |
| 1+ years of age |  |  |  |  |
| Period prevalence | 47,257 | 224,995 | 187,309-272,926 | 75.2, 136.5 |
| Incidence with exclusions | 49,551 | 252,872 | 209,058-308,891 | 74.9, 136.6 |
| Adjusted false positives | 9917 | 37,868 | 31,802-45,596 | 72.9, 123.3 |
| SENSITIVITY ANALYSIS 3† |  |  |  |  |
| < 1 year of age |  |  |  |  |
| Period prevalence | 2050 | 3615 | 2585-5857 | 57.4, 96.8 |
| Incidence with exclusions | 1863 | 3249 | 2341-5192 | 57.4, 96.1 |
| Adjusted false positives | 626 | 769 | 671-963 | 55.1, 86.2 |
| 1+ years of age |  |  |  |  |
| Period prevalence | 42,446 | 171,908 | 119,993-257,414 | 69.2, 129.8 |
| Incidence with exclusions | 46,411 | 207,041 | 143,834-311,111 | 69.4, 130.6 |
| Adjusted false positives | 6541 | 21,678 | 15,685-31,450 | 67.0, 114.5 |

\*AIC = Akaike information criterion and BIC = Bayesian information criterion, for both lower values indicate a comparably better model fit
<sup>†</sup>Sensitivity analysis 1 = including probable iPHIS case reports, Sensitivity analysis 2 = including iPHIS probable and “does not meet” case reports and Labware indeterminate laboratory tests, Sensitivity analysis 3 = removing A37.9 codes with pertussis species unspecified

### Sensitivity analyses

After the addition of probable iPHIS cases, there was little change to estimated abundance (Table 5.1). Including iPHIS probable and DNM cases and indeterminate laboratory tests produced the largest increase in abundance and introduced a considerable amount of variability. While this pattern occurred under all case definitions, the highest estimate of 252,872 (95% CI 209,058-308,891) was obtained under incidence for those one year of age or older. After removing A37.9 codes, abundance estimates increased and decreased without a reliable pattern.

### Estimated sensitivity

Sensitivity estimates are available in Table 2. Results were comparable and generally within a few percentage points with and without multiple episodes per individual (S1 Table). As a result, clustering was determined to have a minimal effect and we reported sensitivity estimates that included multiple episodes per individual. Using all data sources consistently provided the highest sensitivity (Table 2). Labware had the lowest sensitivity while administrative data had the highest for a single source. Under the primary analysis and prevalence definition, the highest sensitivity for those less than one year of age was 69.2% (95% CI 67.7-70.7%), which dropped to 40.6% (95% CI 40.3-40.8%) for the older age group. Incidence sensitivity estimates were similar but slightly lower in comparison, excepting marginally higher estimates for iPHIS and Labware for the younger age group. Overall, using the adjusted false positive definition increased sensitivity, with all sources under the primary analysis producing a sensitivity of 78.5% (95% CI 76.2-80.9%) and 43.1% (95% CI 42.4-43.8%) for the younger and older age groups respectively. However, the largest increase to sensitivity occurred for iPHIS and Labware estimates, although sensitivity for these sources remained low for the older age group.

**Table 2.** Estimated sensitivity by case definition, age group, analysis, and data source.

|  |  | PERIOD PREVALENCE |  |  | INCIDENCE WITH EXCLUSIONS |  |  | ADJUSTED FALSE POSITIVES |  |  |
| --- | --- | --- | --- | --- | --- | --- | --- | --- | --- | --- |
| Analysis | Data source | Sensitivity (%) | (n/N)<sup>§</sup> | 95% CI (%) | Sensitivity (%) | (n/N)<sup>§</sup> | 95% CI (%) | Sensitivity (%) | (n/N)<sup>§</sup> | 95% CI (%) |
| < 1 YEAR OF AGE |  |  |  |  |  |  |  |  |  |  |
| Primary analysis | All data sources | 69.2 | (2636/3810) | 67.7-70.7 | 68.8 | (2118/3078) | 67.2-70.4 | 78.5 | (904/1151) | 76.2-80.9 |
|  | Admin data | 66.0 | (2516/3810) | 64.5-67.5 | 64.8 | (1994/3078) | 63.1-66.5 | 67.8 | (780/1151) | 65.1-70.5 |
|  | Labware | 9.69 | (369/3810) | 8.75-10.6 | 12.0-12.1 | (368-372/3078)* | 10.8-13.2 | 32.0-32.3 | (368-372/1151)* | 29.3-35.0 |
|  | iPHIS | 12.1 | (461/3810) | 11.1-13.1 | 14.8-14.9 | (456-460/3078)* | 13.7-16.2 | 39.6-40.0 | (456-460/1151)* | 36.8-42.8 |
| Sensitivity analysis 1<sup>†</sup> | All data sources | 65.7 | (2648/4030) | 64.2-67.2 | 65.9 | (2133/3235) | 64.3-67.6 | 77.1 | (897/1163) | 74.7-79.5 |
|  | Admin data | 62.4 | (2516/4030) | 60.9-63.9 | 61.6-61.7 | (1993-1997/3235) | 60.1-63.4 | 65.4 | (761/1163) | 63.8-67.0 |
|  | Labware | 9.16 | (369/4030) | 8.27-10.0 | 11.4-11.5 | (368-372/3235)* | 10.3-12.6 | 31.6-32.0 | (368-372/1163)* | 30.1-33.2 |
|  | iPHIS | 11.9 | (481/4030) | 10.9-12.9 | 14.7-14.8 | (476-480/3235)* | 13.5-16.1 | 40.9-41.3 | (476-480/1163)* | 39.6-42.9 |
| Sensitivity analysis 2<sup>†</sup> | All data sources | 29.7 | (2729/9197) | 28.7-30.6 | 28.0 | (2206/7883) | 27.0-29.0 | 41.0 | (1007/2454) | 39.1-43.0 |
|  | Admin data | 27.3-27.4 | (2515-2519/9197)* | 26.4-28.3 | 25.3-25.4 | (1995-1999/7883)* | 24.3-26.3 | 32.6 | (800/2454) | 30.7-34.5 |
|  | Labware | 5.09 | (468/9197) | 4.64-5.54 | 5.95-6.00 | (469-473/7883)* | 5.43-6.52 | 19.1 | (469/2454) | 17.6-20.7 |
|  | iPHIS | 5.96 | (548/9197) | 5.47-6.44 | 6.56 | (517/7883) | 6.01-7.10 | 21.1 | (517/2454) | 19.5-22.7 |
| Sensitivity analysis 3<sup>†</sup> | All data sources | 56.7 | (2050/3615) | 55.1-58.3 | 57.3 | (1863/3249) | 55.6-59.0 | 81.4 | (626/769) | 78.7-84.2 |
|  | Admin data | 50.7 | (1832/3615) | 49.0-52.3 | 50.5 | (1642/3249) | 48.8-52.3 | 52.7 | (405/769) | 49.1-56.2 |
|  | Labware | 10.2-10.3 | (368-372/3615)* | 9.19-11.3 | 11.3-11.4 | (368-372/3249)* | 10.2-12.5 | 47.9-48.4 | (368-372/769)* | 44.3-51.9 |
|  | iPHIS | 12.8 | (461/3615) | 11.7-13.8 | 14.2 | (461/3249) | 13.0-15.4 | 59.9 | (461/769) | 56.5-63.4 |
| 1 + YEARS OF AGE |  |  |  |  |  |  |  |  |  |  |
| Primary analysis | All data sources | 40.6 | (46,295/114,135) | 40.3-40.8 | 36.7 | (48,676/132,528) | 36.5-37.0 | 43.1 | (8823/20,490) | 42.4-43.8 |
|  | Admin data | 39.4 | (44,933/114,135) | 39.1-39.7 | 35.7 | (47,292/132,528) | 35.4-35.9 | 36.3 | (7444/20,490) | 35.7-37.0 |
|  | Labware | 1.43 | (1636/114,135) | 1.36-1.50 | 1.24 | (1637-1641/132,528)* | 1.18-1.30 | 7.99-8.01 | (1637-1641/20,490)* | 7.62-8.38 |
|  | iPHIS | 2.02 | (2307/114,135) | 1.94-2.10 | 1.74 | (2305-2309/132,528)* | 1.67-1.81 | 11.2-11.3 | (2305-2309/20,490)* | 10.8-11.7 |
| Sensitivity analysis 1<sup>†</sup> | All data sources | 45.2 | (46,581/102,954) | 44.9-45.5 | 41.0 | (49,015/119,684) | 40.7-41.2 | 48.6 | (9295/19,120) | 47.9-49.3 |
|  | Admin data | 43.6 | (44,931-44,935/102,954)* | 43.3-43.9 | 39.6 | (47,339/119,684) | 37.9-41.2 | 39.8 | (7619/19,120) | 38.2-41.5 |
|  | Labware | 1.59 | (1636/102,954) | 1.51-1.67 | 1.37 | (1637-1641/119,684)* | 0.98-1.76 | 8.56-8.58 | (1637-1641/19,120)* | 7.62-9.53 |
|  | iPHIS | 2.72 | (2800/102,954) | 2.62-2.82 | 2.34 | (2798-2802/119,684)* | 1.83-2.85 | 14.6-14.7 | (2798-2802/19,120)* | 13.5-15.8 |
| Sensitivity analysis 2<sup>†</sup> | All data sources | 21.0 | (47,257/224,995) | 20.8-21.2 | 19.6 | (49,551/252,872) | 19.4-19.8 | 26.2 | (9917/37,868) | 25.7-26.6 |
|  | Admin data | 20.0 | (44,926/224,995) | 19.8-20.1 | 18.7 | (47,346/252,872) | 18.6-18.9 | 20.4 | (7712/37,868) | 20.0-20.8 |
|  | Labware | 1.03 | (2318/224,995) | 0.99-1.07 | 0.92 | (2318-2322/252,872)* | 0.88-0.96 | 6.12-6.13 | (2318-2322/37,868)* | 5.88-6.37 |
|  | iPHIS | 1.48 | (3332/224,995) | 1.43-1.53 | 1.22 | (3079/252,872) | 1.17-1.26 | 8.13 | (3079/37,868) | 7.86-8.41 |
| Sensitivity analysis 3<sup>†</sup> | All data sources | 24.7 | (42,446/171,908) | 24.5-24.9 | 22.4 | (46,411/207,041) | 22.2-22.6 | 30.2 | (6541/21,678) | 29.6-30.8 |
|  | Admin data | 23.7 | (40,710/171,908) | 23.5-23.9 | 21.6 | (44,657/207,041) | 21.4-21.7 | 22.1 | (4787/21,678) | 20.7-23.5 |
|  | Labware | 0.95 | (1633-1637/171,908)* | 0.90-1.00 | 0.79 | (1633-1637/207,041)* | 0.75-0.83 | 7.53-7.55 | (1633-1637/21,678)* | 6.64-8.44 |
|  | iPHIS | 1.34-1.35 | (2310-2314/171,908)* | 1.29-1.40 | 1.12 | (2311-2315/207,041)* | 1.07-1.16 | 10.7 | (2311-2315/21,678)* | 9.62-11.7 |

\*suppressed for reporting due to low cell size (direct or by inference)
<sup>§</sup>the observed in individual data sources will not sum to the observed in all data sources due to overlap between sources<sup>†</sup>Sensitivity analysis 1 = including probable iPHIS case reports, Sensitivity analysis 2 = including iPHIS probable and “does not meet” case reports and Labware indeterminate laboratory tests,
Sensitivity analysis 3 = removing A37.9 codes with pertussis species unspecified

Any change to sensitivity estimates after the addition of probable iPHIS cases was small, within 5%. After including iPHIS probable and DNM cases and indeterminate laboratory tests, the sensitivity estimates for all sources combined were more similar between age groups. Under prevalence, sensitivity was 29.7% (95% CI 28.7-30.6%) and 21.0% (95% CI 20.8-21.2%) for the younger and older groups respectively. Excluding A37.9 codes under the adjusted false positive definition led to the highest Labware and iPHIS sensitivity estimates for the younger age group, at close to 50% and 60%.

## Discussion

Abundance estimates demonstrated that a high proportion of pertussis cases were missed by all sources. Results were similar when using prevalence and incidence, but after adjusting for physician billing code-only false positives abundance dropped considerably. While this occurred for both age groups, the effect was greater among those one year of age or older. The low estimated PPV of physician billing code-only episodes provides evidence that false positives have been inflating observed counts and abundance estimates, which decreased sensitivity for laboratory and case report data. As a result, the adjusted case definition is the most valid out of those tested, with the described approach useful for improving the utility of capture-recapture methods to epidemiological surveillance. Regardless of the case definition, health administrative data had the highest sensitivity for a single source, with all sources combined producing the best sensitivity. This establishes the value of incorporating health administrative data into pertussis surveillance if accuracy and timeliness limitations are addressed. Laboratory tests had the lowest sensitivity and particularly for the older age group, indicating testing bias is present. Public health case reports had slightly higher sensitivity but displayed a similar pattern. While sensitivity estimates improved for both after adjusting for false positives, this was primarily in the younger age group. Overall, sensitivity was substantially lower for the older group, suggesting ascertainment bias is present.

A 2018 Ontario capture-recapture study used the same data sources but over fewer years. As a result, abundance is not directly comparable as the higher estimates from this study are expected due to the extra years of data. However, sensitivity for all sources with probable case reports included was estimated at 54% and 39% for the younger and older age groups respectively. This is comparable to sensitivity estimates for the older age group in this study after including iPHIS probable cases, but sensitivity was higher for the younger group (∼66%). This could be due to improved recapture in this subgroup using the case definitions or differences in how the 2018 study modelled dependencies [10]. The 2018 study reported considerable uncertainty persisting around physician billing code accuracy and investigated by using different proportions of true positives for all health administrative data, with the lowest at 25%. This dropped abundance estimates by 66% and 73% for the younger and older age groups [10]. We were able to address remaining uncertainty through validation of physician billing code-only episodes. Interestingly, applying the estimated PPV of 8.99% to these episodes reduced abundance estimates similarly to assuming a PPV of 25% for all health administrative data in the 2018 study, by 64% and 84%. The higher latter value indicates a greater proportion of code only-episodes in the older age group, leading to enhanced improvement in recapture once removed. Laboratory test and case report sensitivity considerably improved using this case definition.

A modelling study based on pertussis incidence in southern Ontario from 1993-2004 estimated that five to 33,032 cases remain undetected per reported case depending on age [3]. It has been stated elsewhere that the true number of pertussis cases is at least three times higher than what is reported [2]. To compare, estimates from this study should be calculated using the false positive case definition to avoid inflating underdetection. For case report data, these values are 2.5 and 8.9 for the younger and older age groups under the primary analysis and 4.7 and 12 after including probable and DNM case reports and indeterminate laboratory tests. Using all data sources, 1.3 and 2.3 cases were missed per observed case for the younger and older age groups, which increased to 2.4 and 3.8 with the additional case reports and laboratory tests. The substantially lower upper limit compared to the modelling study is likely due to differences in age groups, pertussis incidence during the respective time periods, and diagnostic testing accuracy, with PCR introduced after 2004. In addition, the estimation approaches differ, with the modelling study using methods with considerable uncertainty as reflected by the wide range of values. While this study used the more conservative Chao’s lower bound estimator, similar results were obtained from Darroch models. Furthermore, a simulation study found that Chao’s methods estimated abundance within 75-82% of the total population size in most complicated scenarios [11]. Regardless, this study’s findings are in line with past estimates of underdetection, with reasonable explanations for remaining differences.

The estimated PPV for physician billing code-only episodes of 8.99% (95% CI 1.59-16.39%) is comparable to the PPV of 13.6% (95% CI 9.28-17.9%) obtained for an OHIP physician billing code algorithm within the EMRPC using the same cohort [16]. The slightly higher PPV in the EMRPC can be explained by using prevalent cases, increasing the likelihood of concordance between codes and cases. Additionally, EMRPC billing codes were only collected from physician offices, potentially decreasing the number of false positives. OHIP pertussis cases billed at homes or long-term care facilities are unlikely to be documented in EMRPC’s primary care patient records. Further contributing to this issue is that visits outside office settings such as walk-in or specialist visits fail to be captured in the EMRPC, leading to about 15% of interactions being missed [24]. In addition, only two thirds of the laboratory tests in OHIP are documented in the EMRPC [24]. Missing any of these data in the EMRPC could artificially decrease the PPV of OHIP billing code-only episodes by increasing the number of false positives, although billings from outside office settings were uncommon in the OHIP database and unlikely to greatly affect validation. The EMRPC study only tested the accuracy of data available in the EMRPC, meaning sensitivity estimates for emergency room visits, hospitalizations, and case report data are not available. However, pertussis laboratory test sensitivity using prevalent cases was reported as 0.64% (95% CI 0.37-1.09%) across all ages [16]. After adjusting for false positives, 8.0% sensitivity (95% CI 7.62-8.38%) was obtained for the older age group (which only excludes infants) using prevalence. This difference can be explained by variation in ages, pertussis classification, and validation methods. Additionally, Labware has more comprehensive coverage, with the EMRPC noted to have decreased laboratory test sensitivity through incomplete documentation [16].

One limitation of this study is that we did not validate physician billing code-only episodes separately for the younger and older age groups. This was to preserve an adequate sample size, with the assumption that the PPV averages out across age groups and this is an appropriate strategy for taking a random sample based on a proportion. In addition, while demonstrating that sensitivity estimates differ by age group, it is unlikely that PPV would vary to the same extent. PPV evaluates the proportion of true positives out of test positives and is primarily affected by prevalence, not testing bias [25]. Although it may appear that younger individuals have a higher risk of pertussis infection, due to testing and ascertainment bias this may not actually be the case [4]. Older cases are hypothesized to be an important source of pertussis, which is evident through older relatives being key sources of transmission to infants [4, 26]. Additionally, in 2019, 62% of reported cases in Ontario occurred in those ten years of age or older [27]. However, it may be of interest to allow the PPV to vary separately by age group in future analyses.

An additional limitation is that we had to remove EMRPC cases without dates during validation. We considered it preferable to avoid introducing misclassification rather than preserving the sample size, and there is no reason to suspect excluded cases are systematically different from the majority of those included. A further limitation of validation is that we were unable to adjust the PPV and sensitivity estimates for clustering under the incidence and false positive definitions due to the low sample size and study methodology respectively [22, 23]. To address this, we compared point estimates and variances to those using a single episode per individual to assess the effect of correlation, with little difference found between PPV estimates. While some sensitivity estimates were statistically significantly different, the absolute difference was small, and this was for the older age group where large sample sizes produced substantial precision. As a result, we concluded that these differences were unlikely to be clinically significant. While it is possible correlation still minorly affected the findings, it is a study strength to be able to report sensitivity estimates under different case definitions. Little validation research has incorporated repeated episodes, which is of interest for acute diseases.

Finally, missing data may impact abundance and sensitivity estimates, but this is an inherent limitation of using secondary data and beyond the scope of this study. This includes failing to collect data on certain cases, such as those that are milder in nature who do not seek health care or may be misdiagnosed. While pertussis infectivity is related to severity, meaning that milder undetected cases are likely less infectious, it is possible that many are still important to transmission. While Labware data only covers pertussis laboratory testing for < 95% of Ontario, we assumed that the remaining tests would not considerably impact abundance estimates [10, 28]. To assess the effect of missing data, we conducted sensitivity analyses incorporating additional data to determine the robustness of capture-recapture abundance estimates. This produced little change to results, except when including probable and DNM cases in addition to indeterminate laboratory tests. Doing so decreased sensitivity and led to greater similarity in capture patterns between the younger and older age groups. This could suggest that older individuals are less likely to meet the confirmed case definition, or that milder infections are frequently missed in younger as well as older individuals. Alternatively, it could be due to increased uncertainty in abundance among both groups under this analysis, stemming from greater uncertainty in true pertussis status.

## Conclusions

This study demonstrated how limited health data accuracy can be accounted for using capture-recapture analyses that employ different pertussis case definitions. The false-positive adjusted case definition helped address past uncertainty in burden estimation and produced results which align with the degree of underdetection reported in the literature; improved capture-recapture estimates can better inform public health policy and prevention. Findings consistently demonstrated that data sources are failing to detect pertussis cases, and particularly laboratory and case report data. The best sensitivity was obtained by using all sources together, with health administrative data having the highest sensitivity for a single source. This indicates the benefit of incorporating real time health administrative data into surveillance if misclassification can be addressed. The results provide further support that pertussis detection differs by age, indicating that ascertainment and testing bias is present in data.

## Data Availability

The datasets from this study are held securely in coded form at ICES. While data sharing agreements with data providers prohibit ICES from making these data publicly available due to privacy concerns and risk of re-identification, access may be granted to those who meet pre-specified criteria for confidential access, available at www.ices.on.ca/DAS.

## Acknowledgements

The authors acknowledge the assistance of Branson Chen and John Wang with data preparation and Arezou Saedi and Mohammad Ali Moinshaghaghi with record abstraction.

## Supporting Information

**S1 Fig. Rules for developing incident case episodes and establishing data re-capture**

**S2 Fig. Heterogeneity graphs from the primary analysis by case definition and age group**

**S1 Table. Estimated sensitivity by age group, analysis, and data source for incidence and adjusted false positive case definitions using a single random episode per person**

**S1 Appendix. Validation of Ontario Health Insurance Plan (OHIP) billing diagnostic code-only case episodes**.

